# Decline in pneumococcal disease in young children during the COVID-19 pandemic associated with suppression of seasonal respiratory viruses, despite persistent pneumococcal carriage: A prospective cohort study

**DOI:** 10.1101/2021.07.29.21261308

**Authors:** Dana Danino, Shalom Ben-Shimol, Bart Adriaan van der Beek, Noga Givon-Lavi, Yonat Shemer Avni, David Greenberg, Daniel M. Weinberger, Ron Dagan

## Abstract

**Background:** Invasive pneumococcal disease (IPD) declined during the COVID-19 pandemic. Previous studies hypothesized that this was due to reduced pneumococcal transmission resulting from non-pharmacological interventions. We used multiple ongoing cohort surveillance projects in children <5 years to test this hypothesis.

**Methods:** The first SARS-CoV-2 cases were detected in February-2020, resulting in a full lockdown, followed by several partial restrictions. Data from ongoing surveillance projects captured the incidence dynamics of community-acquired alveolar pneumonia (CAAP), non-alveolar lower respiratory infections necessitating chest X-rays (NA-LRI), nasopharyngeal pneumococcal carriage in non-respiratory visits, nasopharyngeal respiratory virus detection (by PCR), and nationwide invasive pneumococcal disease (IPD). Monthly rates (January-2020 through February-2021 vs. mean monthly rates 2016-2019 [expected rates]) adjusted for age and ethnicity, were compared.

**Findings:** CAAP and bacteremic pneumococcal pneumonia were strongly reduced (incidence rate ratios, [IRRs] 0η07 and 0η19, respectively); NA-LRI and non-pneumonia IPD were also reduced, with a lesser magnitude (IRRs, 0η46 and 0η42, respectively). In contrast, pneumococcal carriage prevalence was only slightly reduced and density of colonization and pneumococcal serotype distributions were similar to previous years. The pneumococcus-associated disease decline was temporally associated with a full suppression of RSV, influenza viruses, and hMPV, often implicated as co-pathogens with pneumococcus. In contrast, adenovirus, rhinovirus, and parainfluenza activities were within or above expected levels.

**Interpretation:** Reductions in pneumococcal and pneumococcus-associated diseases occurring during the COVID-19 pandemic were not predominantly related to reduced pneumococcal transmission and carriage but were strongly associated with the complete disappearance of specific respiratory viruses.

**Funding:** Partially funded by Pfizer, Inc.

## Introduction

*Streptococcus pneumoniae* (pneumococcus) is a leading cause of acute respiratory and invasive infections in all ages, including young children.^1^ Rates of disease caused by pneumococcus are influenced both by the frequency of exposure to the bacteria (i.e., prevalence of nasopharyngeal carriage among healthy children) and by host susceptibility. Epidemiological, clinical, and experimental evidence suggests that certain viruses can increase the susceptibility of individuals to pneumococcal disease when they are exposed to the bacteria.^1-3^ Since SARS-CoV-2 is a respiratory virus, concerns were raised that COVID-19 patients could have increased susceptibility to pneumococcal infections, in particular pneumonia and invasive pneumococcal diseases (IPD). However, early reports during the COVID-19 pandemic suggested that, in fact, the incidence of IPD had been reduced,^4^ and *S. pneumoniae* only infrequently co-infected patients with COVID-19 disease, including pneumonia.^5^ At the same time, non-pharmacological interventions (NPIs) such as social distancing and travel restrictions surrounding the COVID-19 pandemic were associated with an unexpected global suppression of the activity of several seasonal respiratory viruses, which are often implicated as co-pathogens with pneumococcus. These include respiratory syncytial virus (RSV), influenza viruses, and human metapneumovirus (hMPV).^6,7^ We hypothesized that the reduction in pneumococcal disease in children observed during the COVID-19 pandemic was due mainly to the reduction in the incidence of these seasonal respiratory viruses rather than to reductions in transmission of pneumococcus.

We took advantage of multiple ongoing prospective pediatric cohort surveillance projects in our region to assess the dynamics of various clinical presentations associated with pneumococcus during the COVID-19 pandemic period (January 2020 through February 2021). The patterns of pneumococcal disease during the pandemic were evaluated in comparison to rates during 2016-2019 and to common seasonal respiratory viruses. Uniquely, our data included information on clinical diseases caused by pneumococcus, pneumococcal carriage among healthy children, and the activity of specific respiratory viruses. These data provide an opportunity to understand the factors that influence the incidence of pneumococcal disease in children.

## Methods

### Setting

The Soroka University Medical Center (SUMC) is the only hospital in the Negev district of southern Israel, providing primary health services to the entire population of the region. Over 95% of the children living in the region are served by the SUMC, enabling incidence calculations. Two ethnic populations reside in southern Israel: The Bedouin population, with a similar epidemiology of infectious diseases as lower-middle income countries; and the Jewish population, with an epidemiology resembling higher-income Western countries. Higher rates of respiratory and invasive infections, and pneumococcal carriage were reported among Bedouin than among Jewish children ^8^. In 2020, there were ∼97,400 children under five years old in the Negev district; ∼50% were Bedouin children. (Israel Central Bureau of statistics. https://www.cbs.gov.il/he/publications/doclib/2018/3.%20shnatonvitalstatistics/st03_11x.pdf. Accessed 20 July 2021)

Pneumococcal conjugate vaccines (PCVs) were introduced to the National Immunization Plan (NIP) in 2009 (PCV7) and 2010 (PCV13); since 2013 ∼95% and 90% of children received ≥two and three doses, respectively, with high uptake among both Jewish and Bedouin children.^9^

### COVID-19 in Israel

The first case of SARS-CoV-2 was reported in Israel on February 21^st^ 2020. Three official national lockdown periods, with variable degrees of stringency, were implemented thereafter (supplementary figure 1): 1) April through Mid-May 2020 (including Passover and Ramadan holidays); 2) Mid-September to November 2020 (High Holidays season); and 3) December to early February 2021. The detailed NPIs and restrictions are described in supplementary figure 1. The nationwide daily numbers of new COVID-19 cases increased from less than <50 by the end of the first lockdown to ∼6,000 before the second lockdown, and ∼4,000 before the third lockdown. In mid-September, Israel ranked first globally in the rate of COVID-19 infections per capita, (Israel Ministry of Health. COVID-19 Data Repository of the Israeli Ministry of Health, https://data.gov.il/dataset/covid-19. Accessed 20 July 2021), reaching >8,000 new daily cases by January 2021. However, with increasing vaccination coverage, the rates declined towards mid-February.

### Study design

The study population for pneumonia and virological testing comprised all cohorts of children under five years old born in our region from January 2016 through February 2021. For IPD, we included all children under five years old nationwide. For pneumococcal carriage studies, we included children under three years old in our region. Our data were derived from multiple ongoing, prospective long-standing cohort surveillance programs. Monthly incidence or prevalence rates during 2020-2021 were compared to mean monthly rates during the years 2016 through 2019. Total numbers of cases for each prospective surveillance database and comparison of demographic characteristics, during 2016-2019 and January 2020 through February 2021 are presented in Table 1 (a detailed description of the programs is presented in the supplementary material). In brief, the following outcomes were assessed:

1. *Community-acquired alveolar pneumonia (CAAP) incidence*: Since July 2002, all children under five years old visiting the Pediatric Emergency Room (PER) or hospitalized from whom a chest radiography was obtained were included. CAAP was defined as per the World Health Organization (WHO) Standardization of interpretation of chest radiographs for the diagnosis of pneumonia in children.^10^
2. *Non-alveolar lower respiratory infections* (NA-LRI): All children under five years old visiting the PER or hospitalized for LRIs requiring chest radiography, excluding those with CAAP.^11^
3. *Invasive pneumococcal diseases (IPD)*: This is an ongoing, nationwide prospective, active surveillance, initiated in 1989. All IPD cases in children under five years were reported monthly by local investigators in all 27 medical centers where blood and CSF cultures are obtained, accounting for >95% all IPD cases. A bacteremic pneumonia episode was defined as an illness episode diagnosed as pneumonia by the treating physician during which *S. pneumoniae* was isolated from blood.^12^ A non-pneumonia IPD episode was defined as an IPD episode excluding bacteremic pneumonia.
4. *Nasopharyngeal pneumococcal carriage*:
  a. *Children visiting the SUMC:* Since November 2009, each working day nasopharyngeal cultures have been obtained from the first eight children under five years old seen at the PER or within 48 hours, if hospitalized. Children under three years old without respiratory infections were included in the current analysis. Positive cultures were assessed by semi-quantitative methodology^13^
  b. *Healthy children*: Since 2011, nasopharyngeal cultures have been obtained from healthy children under three years old, presenting for vaccination ^14^.The detailed methodology was previously described.^9^
5. *Nasopharyngeal detection of respiratory viruses in hospitalized children*: Respiratory virus samples were obtained as per clinical indication and sent to the SUMC virology laboratory for detection of RSV, influenza A and B viruses, parainfluenza virus, adenovirus, hMPV and rhinovirus (rhinovirus was added since February 2019), as previously described ^15^ (Supplementary Table 1).
6. *All pediatric PER visits and hospitalizations*: Data were collected from the SUMC computerized records.
7. *Hospital visits for all-cause trauma:* PER visits with trauma diagnosis were identified using the *International Classification of Diseases, Ninth Revision, Clinical Modification* (ICD-9-CM) discharge codes.

**Table 1:** Demographics of the study population.

|  | Total | 2016-2019 | 2020-2021 | p-value* |
| --- | --- | --- | --- | --- |
| Viruses |  |  |  |  |
| Cases | 24,088 | 19,448 | 4,640 |  |
| Age (mean±SD) | 14.2±14.8 | 14.0±14.7 | 14.7±15.2 | 0.005 |
| Ethnicity; n (%) |  |  |  |  |
| Jewish | 8,454 (35.1%) | 12,432 (63.9%) | 3,202 (69.0%) | <0.001 |
| Bedouin | 15,634 (64.9%) | 7,016 (36.1%) | 1,438 (31.0%) |  |
| ER Visits |  |  |  |  |
| Cases | 121,606 | 99,090 | 22,516 |  |
| Age (mean±SD) | 20.9±16.7 | 20.8±16.7 | 21.3±17.0 | <0.001 |
| Ethnicity<sup>†</sup>; n (%) |  |  |  |  |
| Jewish | 55,388 (45.6%) | 45,637 (46.1%) | 9,751 (43.3%) | <0.001 |
| Bedouin | 66,105 (54.4%) | 53,352 (53.9%) | 12,753 (56.7%) |  |
| Trauma |  |  |  |  |
| Cases | 17,585 | 14,165 | 3,420 |  |
| Age (mean±SD) | 28.9±16.1 | 28.9±16.1 | 29.0±16.1 | 0.748 |
| Ethnicity<sup>‡</sup>; n (%) |  |  |  |  |
| Jewish | 8,006 (45.5%) | 6,473 (45.7%) | 1,533 (44.8%) | 0.345 |
| Bedouin | 9,572 (54.5%) | 7,685 (54.3%) | 1,887 (55.2%) |  |
| Carriage |  |  |  |  |
| Cases | 9,330 | 7,922 | 1,408 |  |
| Age (mean±SD) | 11.2±9.3 | 11.3±9.3 | 10.7±9.1 | 0.017 |
| Ethnicity; n (%) |  |  |  |  |
| Jewish | 4,202 (45.0%) | 3,616 (45.6%) | 586 (41.6%) | 0.005 |
| Bedouin | 5,128 (55.0%) | 4,306 (54.4%) | 822 (58.4%) |  |
| CAAP |  |  |  |  |
| Cases | 2,684 | 2,307 | 377 |  |
| Age (mean±SD) | 17.1±14.6 | 17.2±14.6 | 16.9±14.6 | 0.724 |
| Ethnicity; n (%) |  |  |  |  |
| Jewish | 1,020 (38.0%) | 881 (38.2%) | 139 (36.9%) | 0.625 |
| Bedouin | 1,664 (62.0%) | 1,426 (61.8%) | 238 (63.1%) |  |
| Non-CAAP LRI |  |  |  |  |
| Cases | 22,597 | 18,277 | 4,320 |  |
| Age (mean±SD) | 17.3±14.8 | 17.2±14.7 | 17.8±15.3 | 0.019 |
| Ethnicity; n (%) |  |  |  |  |
| Jewish | 9,150 (40.5%) | 7,613 (41.7%) | 1,537 (35.6%) | <0.001 |
| Bedouin | 13,447 (59.5%) | 10,664 (58.3%) | 2,783 (64.4%) |  |
| IPD |  |  |  |  |
| Cases | 704 | 592 | 112 |  |
| Age (mean±SD) | 17.1±12.7 | 17.4±12.7 | 15.8±12.4 | 0.233 |
| Ethnicity; n (%) |  |  |  |  |
| Jewish | 574 (81.5%) | 487 (82.3%) | 87 (77.7%) | 0.251 |
| Non-Jewish | 130 (18.5%) | 105 (17.7%) | 25 (22.3%) |  |

\*p-value 2020-2021 vs. 2016-2019
†113 cases without data on ethnicity
‡7 cases without data on ethnicity

All the above projects, as well as the overall current study were approved by the SUMC Ethics Committee.

### Statistical analysis

Demographic characteristics were compared between the pre-COVID-19 years (2016-2019) and the COVID-19 period (2020-2021) using the Pearson χ2 test or the Student’s t-test, as appropriate. Monthly incidence rates of all-cause PER visits, trauma visits, IPD, CAAP, and NA-LRIs were calculated by age (under two years old, two to four years old) and ethnicity (Bedouin and Jewish children). The monthly proportions of positive pneumococcal nasopharyngeal cultures were calculated out of all samples for children under one year old and two to three year old, except during April-May 2020 (the months of the strict lockdown) due to disruptions in sample collection. The monthly number of detected respiratory viruses were analyzed by age (under two years old, two to four years old) and ethnicity. Rate ratios of incidences and proportions were calculated to compare COVID-19 and pre-COVID-19 periods. All rate ratios were adjusted for ethnicity and age using the Mantel-Haenszel method. The statistical significance threshold was p < 0.05. Data were analyzed using R 4.0.2.^16^

## Results

The first case of COVID-19 was identified on February 21^st^ 2020. All NPIs that followed, along with the dynamics of all new daily nationwide COVID-19 cases, are described in supplementary figure 1. Overall, until February 28, 2021, 777,155 COVID-19 cases were documented.

### The COVID-19 pandemic was associated with reduced rates of IPD

During October 2020 through February 2021, there was a marked reduction in IPD rates *vs*. expected (IRR 0η47; 95% CI: 0η32 - 0η70) (supplementary tables 2, 3; supplementary figure 2). However, while a reduction of 81% was observed for bacteremic pneumonia compared to the pre-COVID period, the observed reduction of non-pneumonia IPD was only 42% (figure 1A-D).

**Figure 1.**
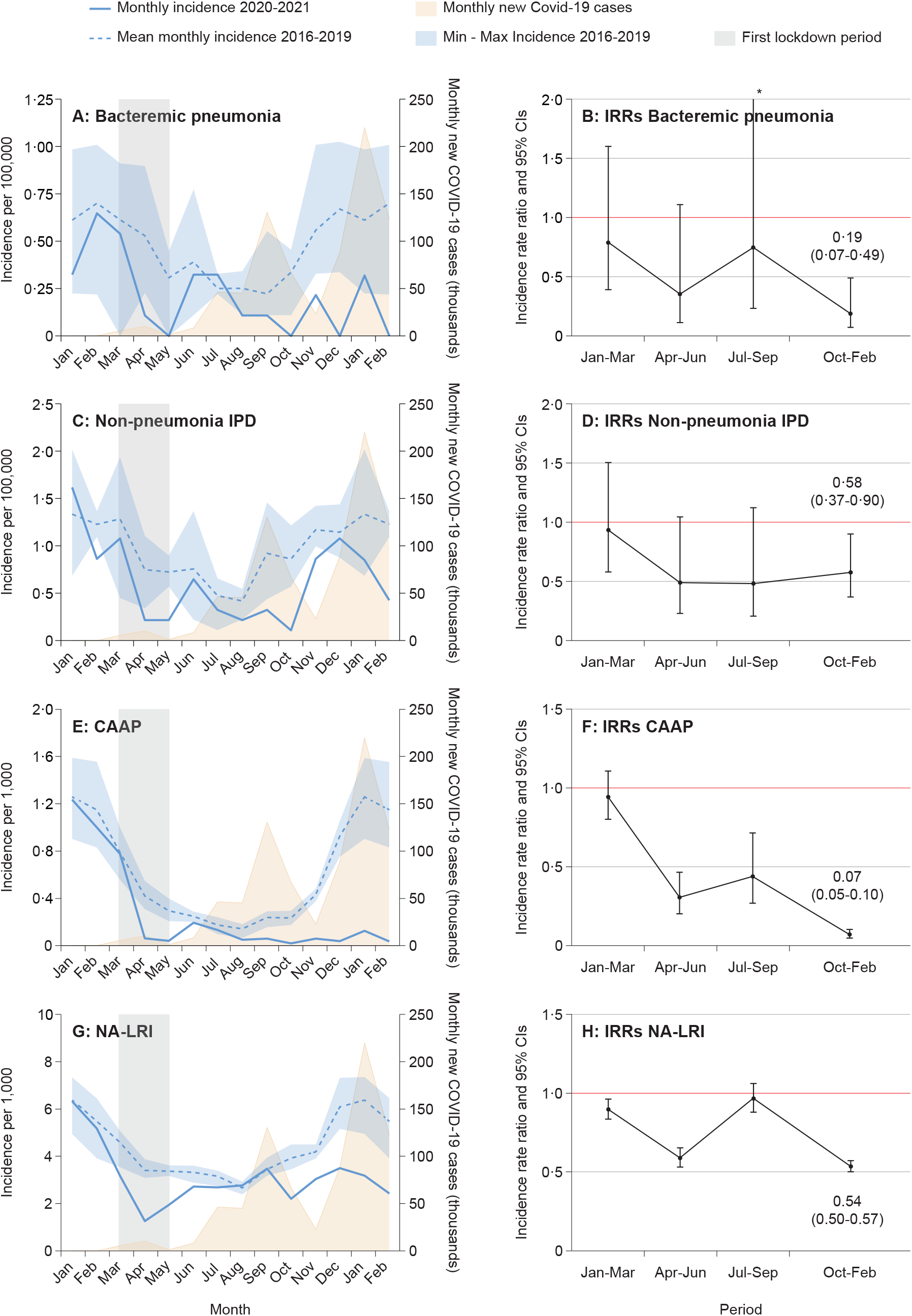
Monthly dynamics of IPD and LRI incidence in children <5 years of age, January 2016 through February 2021, and the numbers of nationally reported new COVID-19 cases (all ages). A- Bacteremic pneumonia C- Non pneumonia IPD E- CAAP G- NA-LRIs Incidence rate ratios (IRRS) by period, January 2029 through February 2021 vs 2016-2019. B- Bacteremic pneumonia D- Non-pneumonia LRI F- CAAP H- NA-LRIs

### The COVID-19 pandemic was associated with reduced CAAP rates, and to a lesser magnitude, reduced NA-LRI rates

Overall LRI rates were reduced during fall and winter 2020-2021 **(supplementary figure 3)**: IRR vs. 2016-2019, 0.47 (95% CI 0.44 – 0.50)

#### Community-acquired alveolar pneumonia

During 2016-2019, a clear seasonality of CAAP, typically thought to be of bacterial etiology, was seen, peaking during December through February and a nadir during June through August (figure 1E, 1F; supplementary tables 2, 4). In the first quarter of 2020, the dynamics of CAAP were similar to those in previous years. During the second quarter, the rates were strongly reduced, coinciding with the strict first lockdown. During the summer months, CAAP rates caught up with the expected seasonal rates, but in October 2020 through February 2021 the rates were extremely low (IRR vs 2016-2019: 0η07; 95% CI 0η05-0η10).

#### Non-alveolar LRIs (NA-LRI)

As with CAAP dynamics, in 2016-2019, NA-LRIs incidence peaked during December through February with a nadir in June through August (figure 1G, 1H; supplementary tables 2, 5). However, the variations between seasons were of a lesser magnitude compared to CAAP. As observed for CAAP, during the first quarter of 2020 dynamics were not affected and the second quarter was affected mainly by the first strict lockdown. However, unlike CAAP rates, following the first lockdown, NA-LRIs rates peaked again to reach the expected seasonal rate in July-September, and the reduction during October 2020 to February 2021 was only half compared to that of CAAP (46%, 95% CI 43-50%; vs 93%, 95% CI 90-95%, respectively), comparing the rates seen in summer and early fall, without the typical peak observed from November through February.

### Nasopharyngeal pneumococcal carriage was not significantly affected during COVID-19 pandemic

During 2016-2019, the mean proportion of children under three years old carrying *S. pneumoniae* was 44η3±1.9%, ranging from 33η8% (August) to 53.8% (December) (figure 2; supplementary table 6). Nasopharyngeal testing was temporarily interrupted during the months of April through May 2020. Although carriage rates were somewhat reduced compared to previous years, during January and February 2021, the rates were very close to those of previous years. Furthermore, using the semi-quantitative method, the mean density was within the ranges of the previous years. Additionally, no notable difference in pneumococcal serotype distribution between the COVID-19 and the pre-COVID period was seen (supplementary table 7).

**Figure 2.**
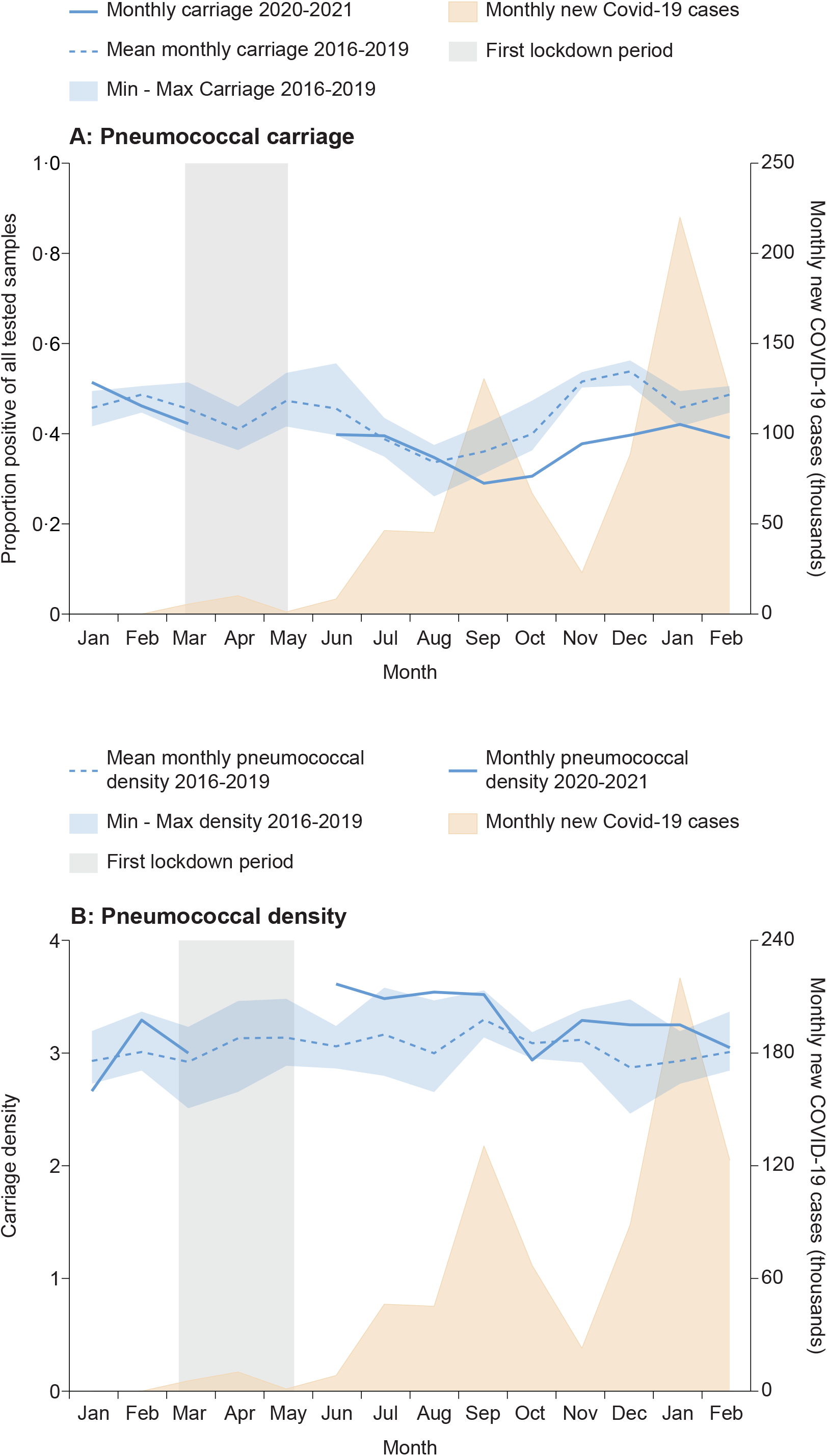
A. Monthly dynamics of proportions of all tested nasopharyngeal swabs that were positive for *S. pneumoniae* in children <3 years of age, and the montly number of newly nationally reported COVID-19 (all ages). B. Dynamics of monthly density of positive nasopharyngeal samples of *S. pneumoniae*, measured by semiquantitative methods, and montly number of newly nationally reported COVID-19 (all ages). min = Minimum max = Maximum Note: During the months of April and May 2000 no samples were collected.

### The COVID-19 pandemic was associated with a marked suppression of RSV, influenza viruses, and hMPV circulation

The detection of respiratory viruses in children during the four years preceding 2020 showed typical dynamics of temperate zone seasonality^17^ (figures 3, 4; supplementary table 8). Apart from rhinovirus, which was common throughout the year, the most prevalent virus was RSV. Fall to early-spring seasonality was seen for RSV, influenza viruses, and hMPV; adenovirus was perennial with some decrease in the summer months; and parainfluenza was lower in winter and spring compared to summer and fall. From January through March 2020, the detection of respiratory viruses was within that expected for the season. During the first (strict) lockdown, very few samples were obtained for viruses, consistent with a strong reduction of all hospital visits. However, thereafter, several important observations were noted: 1) Parainfluenza was undetected from April to November, but sharply increased in December 2020 through February 2021, to levels markedly higher than those seen since 2016; 2) RSV, influenza viruses, and hMPV first decreased during April, to non-detectable levels, as expected seasonally (except for two RSV-detections, in August and September [one in each month]). However, unexpectedly, a complete suppression continued through February 2021; 3) after the first lockdown (March-April 2020), adenovirus and rhinovirus activity resumed to levels observed in previous years (for rhinovirus, only activity since February 2019 was available for comparison).

**Figure 3.**
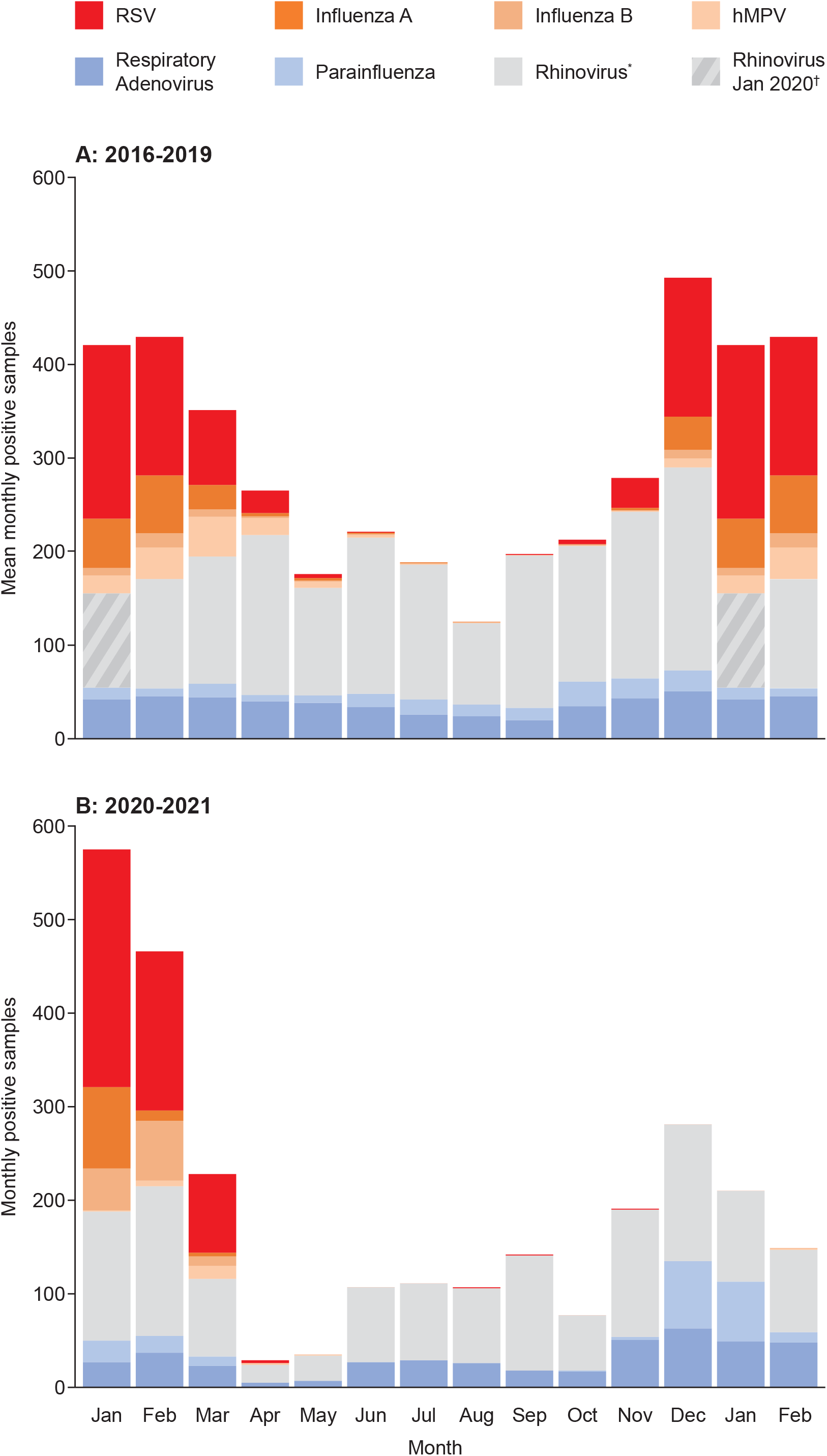
A. Mean monthly virus-positive nasal wash samples in children <5 years of age, by specific virus, 2016 through 2019. *Rhinovirus PCR testing was initiatied in February 2019; thus, for rhinovirus the graph depicts only the year 2019 † Since rhinovirus testing was not done in January 2019, we depicted for January 2019 the result in January 2020 instead B. Monthly virus-positive nasal wash samples in children <5 years of age, by specific virus, January 2020 through February 2021.

**Figure 4.**
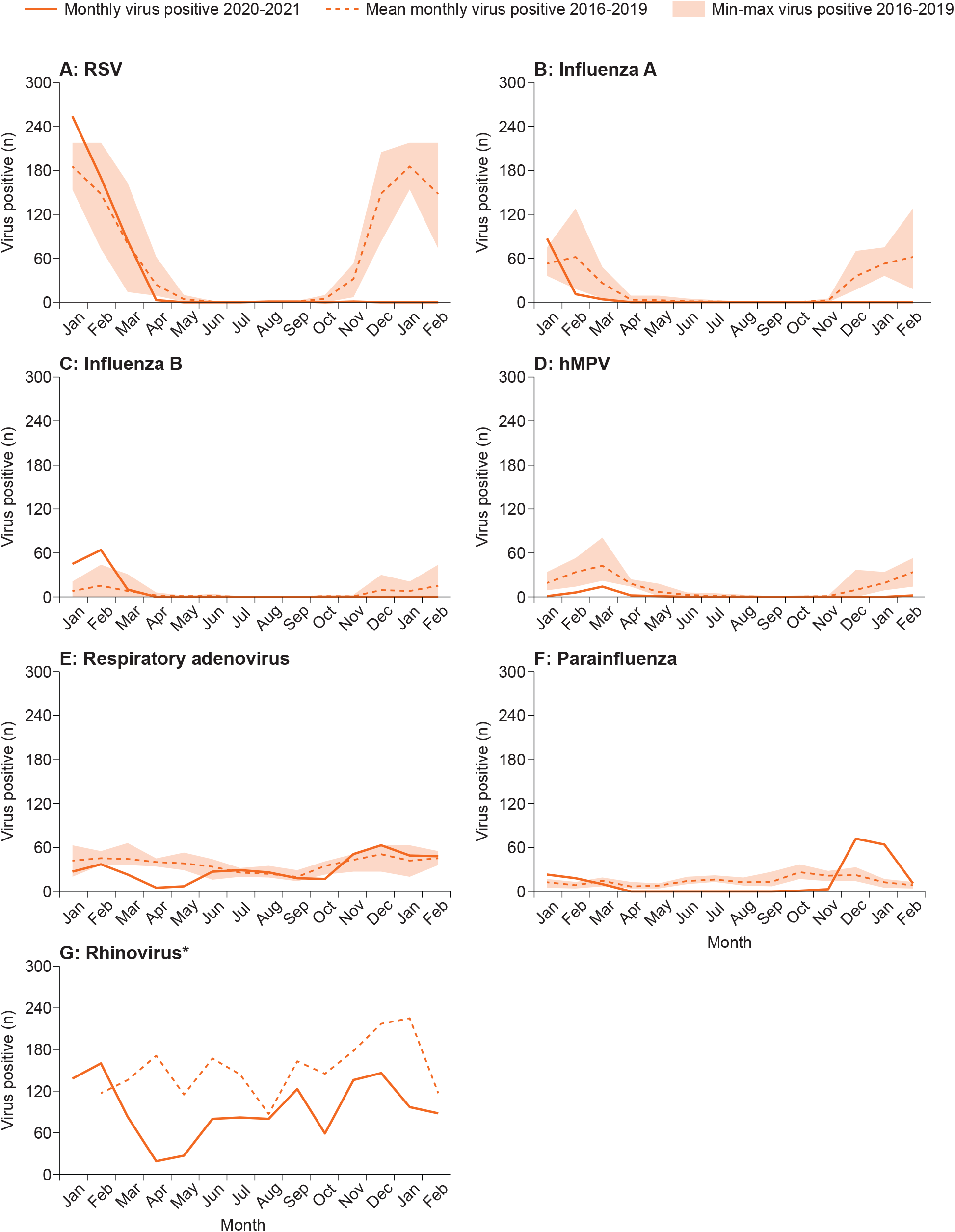
Dynamics of monthly numbers of virus-positive nasal wash samples of specific viruses, January 2016 through February 2021, in children <5 years of age *Rhinovirus PCV usage was initiated in February 2019 min = Minimum max = Maximum

### The dynamics of pneumococcus-associated disease rates were temporally associated with the dynamics of RSV, influenza viruses, and hMPV activity

Throughout the study duration, excluding the period of the first strict lockdown, the dynamics of pneumococcus-associated disease closely followed those of RSV, influenza viruses, and hMPV. This was most striking with bacteremic pneumonia (supplementary figure 4) and CAAP. For NA-LRI and non-pneumonia IPD, although the association was less apparent, the expected typical peak during late fall and winter, coinciding with the expected activity of RSV, influenza viruses, and hMPV, did not occur (exemplified for NA-LRI in supplementary figure 5).

### Hospital visit rates for trauma vs non-trauma

We tested hospitalized visits for trauma, since they are essentially independent of respiratory infections, to examine whether some of the observed reductions in the reporting of LRIs will be related to lower detection rates during the COVID-19 epidemic (supplementary figure 5). As expected, trauma visits peaked in the warm months, in contrast to the non-trauma visits, peaking during fall and winter. For both, a reduction during the first strict lockdown period was seen, but the trauma visit returned to close-to-normal rates, while the overall visits failed to show the expected fall and winter spike.

## Discussion

This prospective cohort study demonstrates a marked reduction of hospital visit rates in southern Israel both for LRI and for pneumococcal disease in children under five years old during the 2020-2021 winter, coinciding with the peak of the COVID-19 pandemic. This reduction was closely associated with an extreme suppression of the circulation of specific respiratory viruses: RSV, influenza viruses, and hMPV. The most dramatic reductions in disease rates were seen in CAAP and bacteremic pneumonia, but NA-LRI and non-pneumonia IPD were also reduced. In contrast, during the same period, pneumococcal nasopharyngeal carriage prevalence was only slightly reduced, and density of colonization and pneumococcal serotype distribution were similar to those observed in previous years. This was a surprising finding because carriage is a prerequisite for pneumococcal disease.

In most published reports, NPIs were suggested to be the cause of the reduction of IPD during the COVID-19 pandemic, assuming that those resulted in the reduction of pneumococcal transmission.^4,18,19^ Our findings lead to the hypothesis that the reduction in pneumococcus-associated disease during fall-winter 2020-2021 resulted from the complete suppression of specific respiratory viruses rather than reduced transmissibility or serotype selection of *S. pneumoniae* in the community. These viruses, especially RSV and influenza are epidemiologically associated with both increased pneumococcal bacteremic pneumonia and non-bacteremic CAAP. ^12,20^ Furthermore, the most frequently detected viruses during IPD and alveolar pneumonia are RSV, followed by influenza viruses and hMPV.^3,21^ The strong association of these viruses with pneumonia might explain why the reduction during fall-winter 2020-2021 was of a higher magnitude in the case of bacteremic pneumonia and CAAP than in non-pneumonia IPD and NA-LRIs.

While, by definition, *S. pneumoniae* is the causative agent for IPD, the extent of pneumococcal involvement in CAAP and NA-LRI is still not fully clarified. This cannot be directly answered, since most frequently, pathogens cannot be isolated in young children and detection of bacteria or viruses in the respiratory tract do not necessarily imply their causative role. However, measuring pneumococcal PCV impact on CAAP and LRI (the “vaccine” probe approach) provides a powerful tool for inference on the likely causative role of *S. pneumoniae* in these clinical outcomes ^22^. Studies from Israel and elsewhere, have demonstrated a ∼50% reduction in CAAP in young children following PCV implementation,^11,23^ thus strongly suggesting an extensive causative role of *S. pneumoniae* in CAAP during the pre-PCV era.

Our NA-LRI cases were severe enough to warrant a chest radiography, but per definition, those with CAAP were excluded. Studies have shown that in such cases, a rate reduction of ≥25% was observed in young children post PCV implementation, including a 34% reduction in our region.^11^ Using a similar logic as that for CAAP, it is plausible that vaccine-serotype pneumococci also played an important role in NA-LRI, although of a lesser magnitude compared to CAAP. The similarity in dynamics between CAAP and IPD, in particular pneumococcal bacteremic pneumonia is therefore not surprising.

Animal and human models have demonstrated that viral infections predispose not only to post-viral pneumococcal diseases, but frequently the true viral-bacterial co-infections.^1^ Using the vaccine probe approach, studies have shown that PCVs could reduce hospitalization for viral infections, including viral pneumonia.^3,24^ Multiple mechanisms by which viruses enhance virulence or invasiveness of *S. pneumoniae* were described, mainly with RSV and influenza virus.^25,26^ It is therefore not surprising that the seasonality of IPD and both bacteremic and non-bacteremic pneumonia are overlapping, especially in regard to CAAP and bacteremic pneumonia.^12^ It is also plausible that the suppression of CAAP and IPD (and bacteremic pneumonia in particular) is deeper than that of NA-LRI diseases for which a lower causative role of *S. pneumoniae* is assumed.

The dynamics of NA-LRIs were similar, but not identical to those of CAAP. After the initial rate reduction due to the strict first lockdown, the rates returned to those seasonally expected, similar to those observed in 2016-2019. In spite of this, the expected late-fall and winter increases did not occur, presumably due to the absence of the abrupt increase of RSV, influenza viruses, and hMPV. In spite of this, the rates were not reduced as much as those of CAAP, but rather showed the same magnitude as those in the summer. The NA-LRI pattern in 2020-21 suggests that viruses such as parainfluenza and adenovirus, and potentially also rhinovirus and others could play a greater relative role for this entity than for CAAP, with a further increase in late-fall and winter rates, related to RSV (the main cause of bronchiolitis in young children) and potentially also to influenza and hMPV.

The main strength of our study is its prospective conduct, using multiple ongoing cohort surveillance programs in the same population, enabling to compare dynamics during the COVID-19 epidemic to those during previous years, on the one hand, and different outcomes, on the other, with appropriate adjustments for time, age and ethnic group. The main weakness of the current study was the inability to determine with certainty the causative agents in the CAAP and NA-LRI cases, a difficulty inherent to all studies on LRI in infants and young children. An additional weakness was that we did not attempt to diagnose all potential viral pathogens including non-COVID-19 corona viruses.

In conclusion, our main message is that unlike common belief, the COVID-19-associated reductions in pneumococcal and pneumococcal-associated diseases in children were not predominantly related to reduced or modified pneumococcal transmission and carriage, but most probably was derived from unprecedented suppression of the activity of specific viruses, capable of increasing the virulence of *S. pneumoniae* in children.

## Supporting information

STROBE CHECKLIST

## Data Availability

We have included all the available data in the manuscript and supplementary file

## Author Contributions

Dana Danino and Ron Dagan contributed to the conception and design of the manuscript, was involved in the acquisition, analysis, and interpretation of data as well as in the drafting and critical review of the manuscript.

Shalom Ben-Shimol contributed to the conception and design of the manuscript, was involved in the acquisition, analysis and interpretation of data, and provided critical review of the manuscript.

Bart Adriaan van der Beek and Noga Givon-Lavi contributed to the conception and design of the manuscript, and to the analysis and interpretation of data, and provided critical review of the manuscript.

Yonat Shemer-Avni has contributed to the acquisition of data and provided critical review of the manuscript.

David Greenberg and Daniel Weinberger contributed to the analysis and interpretation of the data, and provided critical review of the manuscript.

Dana Danino, Adriaan van der Beek, and Noga Givon-Lavi have verified the underlying data.

All authors have approved the final submitted version.

All authors take responsibility for the integrity and accuracy of the data reported.

