## APPENDIX for "Decline in pneumococcal disease in young children during the COVID-19 pandemic associated with suppression of seasonal respiratory viruses, despite persistent pneumococcal carriage: A prospective cohort study"

### Supplementary methods

#### 1) Surveillance of community-acquired alveolar pneumonia (CAAP) and non-alveolar pneumonia LRI (NA-LRI) requiring chest-radiography in children <5 years of age, southern Israel

This is an ongoing, prospective, population-based, active surveillance, initiated in 2002. The current report deals with data from January 2016 through February 2021.
All children <5 years residing in the Negev district, seen at the SUMC PER (either ambulatory or hospitalized within <48 hours from admission) who had CXR examination were included in the analysis.
Clinical practice in the SUMC PER regarding LRI referral, evaluation and the need for CXR examination did not change throughout the study period.
Chest radiographs were analyzed according to the World Health Organization (WHO) Standardization of Interpretation of Chest Radiograph Working Group [1,2]. All chest radiographs were recorded daily and were evaluated separately by the 2 pediatric infectious disease specialists who read, each, all the chest radiographs, independently. Further evaluation was done by a pediatric radiologist who was unaware of the clinical data and of the pediatricians' evaluation. CAAP diagnosis was confirmed by agreement of at least one of the study pediatric infectious disease specialists and the pediatric radiologist [3].

The diagnosis of NA-LRI included all visits of LRI with CXR examination that did not show alveolar infiltrates. This group included both: 1) CXR with readings of infiltrates that were not alveolar (<5% of the group), including linear and patchy densities (interstitial infiltrates) in a lacy pattern involving both lungs, featuring peribronchial thickening and multiple areas of atelectasis; or minor patchy infiltrate that are not of sufficient magnitude to constitute alveolar pneumonia, and small areas of atelectasis [2]; and 2) absence of consolidation, other infiltrates or pleural effusion (but including CXR with hyperinflation, >95% of this group). All NA-LRIs were lumped together, since previous studies showed a very low inter-reader consistency [4].

#### 2) Nationwide Invasive Pneumococcal Diseases (IPD) Surveillance

This has been an ongoing, nationwide, prospective, population-based, active surveillance, initiated in 1989, conducted by the Israeli Pediatric Bacteremia and Meningitis Group (IPBMG) in 27 medical centers routinely obtaining blood cultures from children: All 26 hospitals admitting children and one major outpatient health maintenance organization (HMO, Maccabi Healthcare Services) central laboratory. Less than 1% of blood cultures in all ages and no cerebrospinal fluid (CSF) cultures are obtained outside these centers.
IPD episodes were defined as illness episodes during which *S. pneumoniae* was isolated from blood or CSF. Study population: All children <5 years in Israel. Non-culture diagnoses (polymerase chain reaction [PCR], antigen testing, gram stain or clinical diagnosis only) were excluded. Positive cultures from non-blood or CSF sterile sites (i.e. joint/pleural fluid) were excluded.
Local investigators in each center responded to a monthly distributed questionnaire sent by the principal investigator, located at the Pediatric Infectious Disease Unit (PIDU), the SUMC, which served as the Study headquarters. Several measurements were obtained to ensure data collection; (1) weekly contacts with all 27 laboratories; (2) assuring transportation of all S. pneumoniae isolates to the study headquarters' laboratory; and (3) monthly contact with the local investigators. Completed reports included the following data: Isolate source (blood/CSF), culture date, birthdate, gender, ethnicity (Jewish/non-Jewish), main diagnoses, outcome (mortality) and hospitalization duration. IPD cases reported from each site have been constantly compared to the list of isolates obtained by the reference MOH laboratory. Since July 2009, >95% of the cases have been retrieved [5,6].

#### 3) Pneumococcal nasopharyngeal (NP) carriage surveillance

a) *Children visiting the SUMC:*

This is an ongoing, prospective, population-based, active surveillance, initiated on November 2009. Each working day, nasopharyngeal culture was obtained from the first 4 Jewish and 4 Bedouin children <5 years old, resident of the Negev region, presenting at the PER, whose parents agreed to their enrollment, after obtaining a written informed consent. Children not residing in the Negev region were excluded. In cases where multiple swabs were obtained from a child during the same month, only the first culture was included [7].For the current analysis we used children <3 years of age and excluded those with any diagnosis of respiratory infections. The study was approved by the SUMC Ethics Committee. If <8 children were available, nasopharyngeal carriage evaluation was obtained from randomly selected hospitalized children.

b) *Carriage in Healthy Children*

Since 2011, nasopharyngeal cultures have been obtained from healthy children <3 years old, presenting to the Maternal and Child Health Care Centers in southern Israel for vaccination. A nasopharyngeal swab was obtained after a written informed consent was signed by the parents [8].

Nasopharyngeal samples were obtained using a flexible dacron-tipped swab, introduced through the nostrils and advanced until resistance was found. These swabs were inoculated into modified Stewart transport medium (Medical Wire and Equipment Co., Ltd., Corsham, England) and processed within 16 hours at the SUMC Clinical Microbiology Laboratory. Material from swabs was plated on Columbia agar with 5% sheep blood and 5.0 µg/mL gentamicin, and incubated aerobically at $35^0$C in a $CO_2$-enriched atmosphere for 48h. The presumptive identification of *S. pneumoniae* was based on the presence of α-hemolysis and the inhibition by optochin, and the identity of the bacteria present was confirmed by a positive slide agglutination test result (Phadebact; Pharmacia Diagnostics). One *S. pneumoniae* colony per plate was then subcultured, harvested, and kept frozen at $-70^0$C for further testing. Density analysis was performed at the time of initial sample processing. A semiquantitative plating method was used and graded 0 (negative) to 4 (growth in all plate quadrants). The NP swabs were plated by rolling the swab over one quarter of the plate, and streaking the sample onto 4 quadrants using a sterile loop. Growth was termed 1+, 2+, 3+, or 4+ when colonies were seen in 1, 2, 3, or 4 quadrants, respectively [9].

#### 4) Nasopharyngeal detection of respiratory viruses

Nasopharyngeal wash for respiratory viruses: RSV, influenza A and B viruses, parainfluenza virus, adenovirus, human metapneumovirus (hMPV) and rhinovirus (added in 2019), were taken from hospitalized children according to the treating physician request. Nasopharyngeal specimens, obtained using 2 to 5 mL, of 0.9% saline solution with a mucus extractor kit (Maersk Medical A/S, Lynge, Denmark), were sent to the SUMC Clinical Virology Laboratory within 6 hours. The specimens were diluted with 4mL of RPMI culture medium (Kibbutz Beit Haemek, Israel). Nucleic acid extraction was performed using NucliSENS easy MAG (bioMérieux, Marcy l'Etoile, France), according to the manufacturer's instructions. The sets of primers and probes used to detect the viruses by multiplex hydrolysis probes-based real-time polymerase chain reaction (mqRT-PCR) are shown in **Supplementary Table 1**. Since 2018 viruses detection and differentiation was done using multiplex real time PCR (Seeplex RV7 Detection kt, Seegene Korea) [10].

**Supplementary Table 1: Primers for Respiratory Viruses used in pentaplex real-time RT-PCR assays**

| Pathogen | Primer/Probe | Target gene | Reference or source |
|---|---|---|---|
| Respiratory Synclinal Virus (RSV) | GCCAAAAAATTGTTTCCACAATA<br>TCTTCATCACCATACTTTTCTGTTA<br>ROX-TCAGTAGTAGACCATGTGAATTCCCTGCA-BBQ | L | Modified from reference 9 |
| Influenza A Virus | GGACCTCCACTTACTCCAAAACAGAAAC<br>GTAAGGCTTGCATGAATGTTATTTGCTC<br>YAK-AA_GTTT_GAA_GARATMA_GAT_GGCT-BBQ | NS | Modified from reference 9 |
| Influenza B Virus | ATCGGATCCTCAACTCACTCTT<br>TGACCAAATTGGGATAAGACTC<br>FAM/YAK-CTCGAATTGGCTTTGRATGTCCTTCAT-BBQ | NS | |
| Parainfluenza virus type 2 | ATCCAATCGATACTCGGAGGT<br>TCTGGTTGTTTGGTTGTCCA<br>Cyan500-TGATGGTGAGGACAGAATTGACAAC-BBQ | N | |
| Parainfluenza virus type 3 | AAGATCTACAAGTTGGCAYAGCAA<br>AATGTCCCCATGGACATTCAT<br>ROX-TTCCTGGTCTTGATAGCACATTATGCCA-BBQ | HN | |
| Adenovirus set I | ATGACTTTTGAGGTGGATCCCATGGA<br>GCCGAGAAGGGCGTGCGCAGGTA<br>Cyan500-AGCCCACCCTKC_T_T_TA_T-BBQ | H | |
| Adenovirus set II | GCCCCAGTGGTCTTACATGCACATC<br>GCCACGGTGGGGTTTCTAAACTT<br>Cyan500-TCGGAGTACCTGAGCCCGGGTCTGGTGCA-BBQ | H | 10 |
| Human Metapneumovirus | AACCGTGTACTAAGTGATGCACTC<br>CATTGTTTGACCGGCCCCATAA<br>FAM-CTTTGCCATACTCAATGAACAAACT-BBQ | Np | 11 |
| Rhinovirus | TGGACAGGGTGTGAAGAGC<br>CAAAGTAGTCGGTCCCATCC<br>FAM-TCCTCCGGCCCCTGAATG-BHQ1 | 5_ UTR | 12 |

HN, Hemagglutinin-neuraminidase; NS, nonstructural protein; UTR, untranslated region; H, hexon protein; N, nucleocapsid protein; Np, nucleocapsid phosphoprotein; FAM, 6-carboxyfluorescein; YAK, Yakima; BBQ, BlackBerry Quencher; ROX, carboxy-X-rhodamine; BHQ1, Black Hole Quencher 1.

**Supplementary table 2: IRRs 2020-21 *vs.* 2016-19 per quarter; children < 5 yrs.**

| | Jan-Mar | Apr-Jun | Jul-Sep | Oct-Feb |
|---|---|---|---|---|
| CAAP | 0·94 (0.80 - 1.11) | 0·31 (0.20 - 0.47) | 0·44 (0.27 - 0.71) | 0·07 (0.05 - 0·10) |
| Non-CAAP LRI | 0·90 (0·83 - 0·96) | 0·59 (0·53 - 0·65) | 0·97 (0·88 - 1·06) | 0·54 (0·50 - 0·57) |
| Bacteremic Pneumonia | 0·79 (0·39 - 1·60) | 0·35 (0·11 - 1·11) | 0·75 (0·23 - 2·40) | 0·19 (0·07 - 0·49) |
| Non-pneumonia IPD | 0·93 (0·58 - 1·50) | 0·49 (0·23 - 1·04) | 0·48 (0·21 - 1·12) | 0·58 (0·37 - 0·90) |
| Trauma ER visits | 0·76 (0·68 - 0·84) | 0·67 (0·61 - 0·74) | 0·78 (0·71 - 0·85) | 0·85 (0·79 - 0·92) |
| Non-trauma ER visits | 0·87 (0·84 - 0·90) | 0·56 (0·54 - 0·59) | 0·83 (0·80 - 0·86) | 0·60 (0.58 - 0·62) |
| Carriage* | 1·01 (0·87 - 1·16) | ND | 0·97 (0·81 - 1·16) | 0·72 (0·64 - 0·81) |

*PRRs in children <3 yrs.

**Supplementary table 3: IPD cases children <5 yrs. in Israel per month per study year**

| | 2016 | 2017 | 2018 | 2019 | 2020 | 2021 |
|---|---|---|---|---|---|---|
| Jan | 12 | 20 | 14 | 24 | 18 | 12 |
| Feb | 20 | 20 | 17 | 12 | 14 | 4 |
| Mar | 25 | 4 | 20 | 19 | 15 | |
| Apr | 5 | 13 | 15 | 13 | 3 | |
| May | 8 | 12 | 6 | 11 | 2 | |
| Jun | 17 | 6 | 9 | 9 | 9 | |
| Jul | 7 | 7 | 3 | 9 | 6 | |
| Aug | 7 | 3 | 7 | 7 | 3 | |
| Sep | 7 | 14 | 15 | 5 | 4 | |
| Oct | 9 | 10 | 15 | 9 | 1 | |
| Nov | 15 | 18 | 13 | 16 | 10 | |
| Dec | 21 | 11 | 17 | 16 | 11 | |
| Yearly | 153 | 138 | 151 | 150 | 96 | NA |

**Supplementary table 4: CAAP incidence per 1,000 children <5 yrs. per month per study year**

| | 2016 | 2017 | 2018 | 2019 | 2020 | 2021 |
|---|---|---|---|---|---|---|
| Jan | 1·29 | 1·61 | 0·92 | 1·29 | 1·24 | 0·13 |
| Feb | 0·85 | 1·58 | 1·16 | 1·07 | 1·01 | 0·04 |
| Mar | 0·69 | 0·92 | 0·57 | 1·01 | 0·78 | |
| Apr | 0·56 | 0·34 | 0·32 | 0·50 | 0·06 | |
| May | 0·36 | 0·21 | 0·4 | 0·22 | 0·04 | |
| Jun | 0·27 | 0·29 | 0·21 | 0·24 | 0·20 | |
| Jul | 0·19 | 0·10 | 0·17 | 0·24 | 0·13 | |
| Aug | 0·19 | 0·07 | 0·16 | 0·16 | 0·05 | |
| Sep | 0·16 | 0·25 | 0·29 | 0·25 | 0·06 | |
| Oct | 0·30 | 0·25 | 0·23 | 0·18 | 0·02 | |
| Nov | 0·45 | 0·38 | 0·43 | 0·49 | 0·06 | |
| Dec | 0·95 | 1·07 | 0·76 | 0·97 | 0·04 | |
| Yearly | 6·26 | 7·07 | 5·62 | 6·62 | 3·70 | NA |

**Supplementary table 5: Non-CAAP LRI incidence per 1,000 children <5 yrs. per month per study year**

| | 2016 | 2017 | 2018 | 2019 | 2020 | 2021 |
|---|---|---|---|---|---|---|
| Jan | 5·04 | 6·77 | 6·52 | 7·4 | 6·37 | 3·19 |
| Feb | 3·98 | 6·02 | 6·52 | 5·58 | 5·21 | 2·44 |
| Mar | 3·60 | 4·66 | 5·09 | 5·11 | 3·19 | |
| Apr | 3·32 | 3·12 | 3·42 | 3·92 | 1·27 | |
| May | 3·47 | 3·36 | 3·66 | 3·19 | 1·96 | |
| Jun | 2·93 | 3·39 | 3·48 | 3·64 | 2·74 | |
| Jul | 2·79 | 3·13 | 3·44 | 3·39 | 2·71 | |
| Aug | 2·99 | 2·42 | 2·92 | 2·47 | 2·79 | |
| Sep | 3·42 | 3·37 | 3·99 | 3·21 | 3·50 | |
| Oct | 3·88 | 3·50 | 4·55 | 3·93 | 2·22 | |
| Nov | 3·99 | 4·55 | 4·32 | 4·11 | 3·06 | |
| Dec | 5·26 | 7·40 | 5·52 | 6·46 | 3·52 | |
| Yearly | 44·68 | 51·69 | 53·44 | 52·37 | 38·54 | NA |

**Supplementary table 6: Pnc carriage rates in children <3 yrs.**

| | 2016 | 2017 | 2018 | 2019 | 2020* | 2021 |
|---|---|---|---|---|---|---|
| Jan | 0·42 | 0·48 | 0·49 | 0·44 | 0·51 | 0·42 |
| Feb | 0·51 | 0·50 | 0·50 | 0·45 | 0·46 | 0·39 |
| Mar | 0·40 | 0·44 | 0·51 | 0·47 | 0·42 | |
| Apr | 0·46 | 0·43 | 0·36 | 0·39 | - | |
| May | 0·47 | 0·53 | 0·47 | 0·42 | - | |
| Jun | 0·56 | 0·40 | 0·42 | 0·45 | 0·40 | |
| Jul | 0·43 | 0·38 | 0·35 | 0·38 | 0·40 | |
| Aug | 0·38 | 0·34 | 0·37 | 0·26 | 0·35 | |
| Sep | 0·42 | 0·31 | 0·35 | 0·36 | 0·29 | |
| Oct | 0·47 | 0·36 | 0·39 | 0·37 | 0·31 | |
| Nov | 0·51 | 0·50 | 0·51 | 0·54 | 0·38 | |
| Dec | 0·55 | 0·51 | 0·53 | 0·56 | 0·40 | |
| Yearly | 0·47 | 0·43 | 0·45 | 0·42 | 0·39 | NA |

*No data collection during April and May 2020

**Supplementary table 7: Pnc serotype distribution per study year**

| Serotype | 2016 | 2017 | 2018 | 2019 | 2016-2019 | 2020-2021* |
|---|---|---|---|---|---|---|
| 15B/C | 128 (11·43%) | 142 (13·41%) | 94 (11·84%) | 83 (12·97%) | 447 (12·37%) | 50 (8·76%) |
| 23B | 47 (4·20%) | 65 (6·14%) | 65 (8·19%) | 42 (6·56%) | 219 (6·06%) | 44 (7·71%) |
| 11A | 55 (4·91%) | 60 (5·67%) | 54 (6·80%) | 42 (6·56%) | 211 (5·84%) | 40 (7·01%) |
| 16F | 55 (4·91%) | 55 (5·19%) | 43 (5·42%) | 36 (5·63%) | 189 (5·23%) | 43 (7·53%) |
| 35B | 53 (4·73%) | 56 (5·29%) | 40 (5·04%) | 35 (5·47%) | 184 (5·09%) | 25 (4·38%) |
| 21 | 40 (3·57%) | 61 (5·76%) | 30 (3·78%) | 32 (5·00%) | 163 (4·51%) | 27 (4·73%) |
| 15A | 44 (3·93%) | 35 (3·31%) | 46 (5·79%) | 26 (4·06%) | 151 (4·18%) | 30 (5·25%) |
| 9N | 47 (4·20%) | 36 (3·40%) | 39 (4·91%) | 20 (3·13%) | 142 (3·93%) | 25 (4·38%) |
| 17F | 48 (4·29%) | 32 (3·02%) | 33 (4·16%) | 26 (4·06%) | 139 (3·85%) | 14 (2·45%) |
| 23A | 45 (4·02%) | 35 (3·31%) | 23 (2·90%) | 23 (3·59%) | 126 (3·49%) | 24 (4·20%) |
| 10A | 29 (2·59%) | 34 (3·21%) | 26 (3·27%) | 17 (2·66%) | 106 (2·93%) | 14 (2·45%) |
| 10B | 30 (2·68%) | 20 (1·89%) | 21 (2·64%) | 24 (3·75%) | 95 (2·63%) | 11 (1·93%) |
| 19F | 24 (2·14%) | 29 (2·74%) | 16 (2·02%) | 25 (3·91%) | 94 (2·60%) | 19 (3·33%) |
| 34 | 29 (2·59%) | 28 (2·64%) | 15 (1·89%) | 17 (2·66%) | 89 (2·46%) | 29 (5·08%) |
| 19A | 30 (2·68%) | 24 (2·27%) | 18 (2·27%) | 6 (0·94%) | 78 (2·16%) | 10 (1·75%) |
| 22F | 36 (3·21%) | 22 (2·08%) | 13 (1·64%) | 4 (0·63%) | 75 (2·08%) | 9 (1·58%) |
| 31 | 18 (1·61%) | 20 (1·89%) | 14 (1·76%) | 10 (1·56%) | 62 (1·72%) | 7 (1·23%) |
| 24F | 17 (1·52%) | 14 (1·32%) | 21 (2·64%) | 8 (1·25%) | 60 (1·66%) | 8 (1·40%) |
| 14 | 28 (2·50%) | 17 (1·61%) | 13 (1·64%) | 3 (0·47%) | 61 (1·69%) | 5 (0·88%) |
| 13 | 15 (1·34%) | 12 (1·13%) | 18 (2·27%) | 9 (1·41%) | 54 (1·49%) | 8 (1·40%) |
| 33F | 12 (1·07%) | 13 (1·23%) | 12 (1·51%) | 15 (2·34%) | 52 (1·44%) | 7 (1·23%) |
| 7B | 16 (1·43%) | 13 (1·23%) | 9 (1·13%) | 12 (1·88%) | 50 (1·38%) | 6 (1·05%) |
| 19B | 10 (0·89%) | 9 (0·85%) | 12 (1·51%) | 15 (2·34%) | 46 (1·27%) | 10 (1·75%) |
| 38 | 19 (1·70%) | 12 (1·13%) | 8 (1·01%) | 9 (1·41%) | 48 (1·33%) | 4 (0·70%) |
| 6C | 13 (1·16%) | 11 (1·04%) | 8 (1·01%) | 11 (1·72%) | 43 (1·19%) | 16 (2·80%) |
| 12F | 13 (1·16%) | 13 (1·23%) | 8 (1·01%) | 7 (1·09%) | 41 (1·13%) | 1 (0·18%) |
| 23F | 14 (1·25%) | 9 (0·85%) | 4 (0·50%) | 2 (0·31%) | 29 (0·80%) | 4 (0·70%) |
| 3 | 9 (0·80%) | 7 (0·66%) | 7 (0·88%) | 3 (0·47%) | 26 (0·72%) | 3 (0·53%) |
| 10F | 11 (0·98%) | 10 (0·94%) | 4 (0·50%) | 0 (0·00%) | 25 (0·69%) | 4 (0·70%) |
| 35F | 13 (1·16%) | 4 (0·38%) | 5 (0·63%) | 1 (0·16%) | 23 (0·64%) | 4 (0·70%) |
| 20 | 7 (0·63%) | 4 (0·38%) | 5 (0·63%) | 4 (0·63%) | 20 (0·55%) | 6 (1·05%) |
| 18C | 6 (0·54%) | 8 (0·76%) | 0 (0·00%) | 2 (0·31%) | 16 (0·44%) | 1 (0·18%) |
| 35A | 4 (0·36%) | 5 (0·47%) | 2 (0·25%) | 1 (0·16%) | 12 (0·33%) | 2 (0·35%) |
| 33A | 0 (0·00%) | 9 (0·85%) | 0 (0·00%) | 1 (0·16%) | 10 (0·28%) | 0 (0·00%) |
| 6A | 4 (0·36%) | 6 (0·57%) | 1 (0·13%) | 0 (0·00%) | 11 (0·30%) | 1 (0·18%) |
| 8 | 4 (0·36%) | 4 (0·38%) | 1 (0·13%) | 1 (0·16%) | 10 (0·28%) | 2 (0·35%) |
| 11B | 2 (0·18%) | 0 (0·00%) | 3 (0·38%) | 3 (0·47%) | 8 (0·22%) | 0 (0·00%) |
| 28A | 3 (0·27%) | 2 (0·19%) | 1 (0·13%) | 1 (0·16%) | 7 (0·19%) | 2 (0·35%) |
| 2 | 2 (0·18%) | 0 (0·00%) | 2 (0·25%) | 1 (0·16%) | 5 (0·14%) | 0 (0·00%) |
| 33B | 2 (0·18%) | 0 (0·00%) | 2 (0·25%) | 1 (0·16%) | 5 (0·14%) | 1 (0·18%) |
| 35B/D | 1 (0·09%) | 2 (0·19%) | 1 (0·13%) | 1 (0·16%) | 5 (0·14%) | 1 (0·18%) |
| 7C | 0 (0·00%) | 3 (0·28%) | 1 (0·13%) | 1 (0·16%) | 5 (0·14%) | 1 (0·18%) |
| 27 | 1 (0·09%) | 3 (0·28%) | 0 (0·00%) | 0 (0·00%) | 4 (0·11%) | 0 (0·00%) |
| 6B | 0 (0·00%) | 3 (0·28%) | 1 (0·13%) | 0 (0·00%) | 4 (0·11%) | 0 (0·00%) |
| 18A | 2 (0·18%) | 1 (0·09%) | 0 (0·00%) | 1 (0·16%) | 4 (0·11%) | 1 (0·18%) |
| 35C | 1 (0·09%) | 1 (0·09%) | 1 (0·13%) | 0 (0·00%) | 3 (0·08%) | 0 (0·00%) |
| 9V | 2 (0·18%) | 0 (0·00%) | 1 (0·13%) | 0 (0·00%) | 3 (0·08%) | 0 (0·00%) |
| 24B | 1 (0·09%) | 0 (0·00%) | 0 (0·00%) | 1 (0·16%) | 2 (0·06%) | 0 (0·00%) |
| 1 | 0 (0·00%) | 0 (0·00%) | 1 (0·13%) | 0 (0·00%) | 1 (0·03%) | 1 (0·18%) |
| 22A | 0 (0·00%) | 1 (0·09%) | 0 (0·00%) | 0 (0·00%) | 1 (0·03%) | 0 (0·00%) |
| 25A | 0 (0·00%) | 0 (0·00%) | 1 (0·13%) | 0 (0·00%) | 1 (0·03%) | 1 (0·18%) |
| 29 | 1 (0·09%) | 0 (0·00%) | 0 (0·00%) | 0 (0·00%) | 1 (0·03%) | 0 (0·00%) |
| 35D | 0 (0·00%) | 0 (0·00%) | 1 (0·13%) | 0 (0·00%) | 1 (0·03%) | 0 (0·00%) |
| 36 | 1 (0·09%) | 0 (0·00%) | 0 (0·00%) | 0 (0·00%) | 1 (0·03%) | 0 (0·00%) |
| 37 | 0 (0·00%) | 0 (0·00%) | 0 (0·00%) | 1 (0·16%) | 1 (0·03%) | 0 (0·00%) |
| 4 | 0 (0·00%) | 1 (0·09%) | 0 (0·00%) | 0 (0·00%) | 1 (0·03%) | 0 (0·00%) |
| OMNI NEG | 128 (11·43%) | 108 (10·20%) | 50 (6·30%) | 57 (8·91%) | 343 (9·49%) | 50 (8.76%) |
| Total | 1,120 (100%) | 1,059 (100%) | 794 (100%) | 640 (100%) | 3,613 (100%) | 571 (100%) |

*January 2020 through February 2021 with no data collection during April and May 2020

**Supplementary table 8: Detected viruses per month**

| Year | Month | RSV | Influenza A | Influenza B | hMPV | Respiratory Adenovirus | Paraflu | Rhinovirus* | Total |
|---|---|---|---|---|---|---|---|---|---|
| 2016 | Jan | 154 | 49 | 11 | 15 | 49 | 5 | - | 283 |
| | Feb | 73 | 24 | 44 | 14 | 36 | 4 | - | 195 |
| | Mar | 14 | 7 | 31 | 22 | 37 | 19 | - | 130 |
| | Apr | 10 | 1 | 6 | 14 | 45 | 13 | - | 89 |
| | May | 5 | 1 | 2 | 6 | 39 | 10 | - | 63 |
| | Jun | 1 | 0 | 0 | 5 | 39 | 14 | - | 59 |
| | Jul | 0 | 0 | 1 | 0 | 32 | 13 | - | 46 |
| | Aug | 0 | 0 | 0 | 3 | 35 | 13 | - | 51 |
| | Sep | 2 | 1 | 0 | 1 | 22 | 9 | - | 35 |
| | Oct | 4 | 1 | 0 | 0 | 40 | 17 | - | 62 |
| | Nov | 31 | 2 | 0 | 1 | 50 | 21 | - | 105 |
| | Dec | 106 | 32 | 0 | 0 | 51 | 20 | - | 209 |
| 2017 | Jan | 159 | 51 | 0 | 18 | 63 | 17 | - | 308 |
| | Feb | 119 | 18 | 0 | 53 | 55 | 9 | - | 254 |
| | Mar | 53 | 5 | 0 | 81 | 66 | 17 | - | 222 |
| | Apr | 9 | 1 | 1 | 24 | 34 | 5 | - | 74 |
| | May | 1 | 1 | 1 | 1 | 53 | 11 | - | 68 |
| | Jun | 0 | 0 | 4 | 0 | 44 | 13 | - | 61 |
| | Jul | 0 | 1 | 0 | 0 | 28 | 16 | - | 45 |
| | Aug | 0 | 0 | 1 | 0 | 19 | 10 | - | 30 |
| | Sep | 2 | 0 | 0 | 0 | 14 | 10 | - | 26 |
| | Oct | 5 | 0 | 3 | 0 | 23 | 22 | - | 53 |
| | Nov | 37 | 0 | 3 | 0 | 44 | 14 | - | 98 |
| | Dec | 200 | 17 | 30 | 0 | 63 | 14 | - | 324 |
| 2018 | Jan | 218 | 36 | 21 | 9 | 36 | 16 | - | 336 |
| | Feb | 218 | 77 | 17 | 34 | 39 | 13 | - | 398 |
| | Mar | 90 | 48 | 1 | 44 | 36 | 15 | - | 234 |
| | Apr | 15 | 9 | 1 | 16 | 36 | 4 | - | 81 |
| | May | 1 | 9 | 0 | 18 | 29 | 7 | - | 64 |
| | Jun | 1 | 5 | 0 | 6 | 16 | 10 | - | 38 |
| | Jul | 0 | 3 | 0 | 5 | 23 | 14 | - | 45 |
| | Aug | 0 | 1 | 0 | 0 | 19 | 8 | - | 28 |
| | Sep | 0 | 0 | 0 | 0 | 14 | 7 | - | 21 |
| | Oct | 0 | 1 | 0 | 2 | 35 | 29 | - | 67 |
| | Nov | 7 | 3 | 0 | 2 | 27 | 23 | - | 62 |
| | Dec | 83 | 23 | 0 | 37 | 27 | 22 | - | 192 |
| 2019 | Jan | 211 | 75 | 0 | 34 | 20 | 12 | - | 352 |
| | Feb | 182 | 128 | 0 | 34 | 51 | 8 | 116 | 519 |
| | Mar | 163 | 44 | 0 | 23 | 38 | 7 | 134 | 409 |
| | Apr | 62 | 3 | 0 | 18 | 45 | 5 | 171 | 304 |
| | May | 10 | 0 | 1 | 2 | 32 | 4 | 114 | 163 |
| | Jun | 2 | 0 | 1 | 0 | 36 | 20 | 165 | 224 |
| | Jul | 0 | 0 | 0 | 0 | 20 | 22 | 143 | 185 |
| | Aug | 0 | 1 | 0 | 0 | 23 | 19 | 84 | 127 |
| | Sep | 0 | 0 | 0 | 0 | 29 | 27 | 161 | 217 |
| | Oct | 10 | 0 | 1 | 0 | 41 | 37 | 145 | 234 |
| | Nov | 53 | 5 | 0 | 1 | 51 | 28 | 177 | 315 |
| | Dec | 205 | 70 | 7 | 1 | 62 | 33 | 211 | 589 |
| 2020 | Jan | 254 | 87 | 45 | 1 | 27 | 23 | 138 | 575 |
| | Feb | 170 | 11 | 64 | 6 | 37 | 18 | 160 | 466 |
| | Mar | 84 | 4 | 10 | 14 | 23 | 10 | 83 | 228 |
| | Apr | 3 | 0 | 0 | 2 | 5 | 0 | 19 | 29 |
| | May | 0 | 0 | 0 | 1 | 7 | 0 | 27 | 35 |
| | Jun | 0 | 0 | 0 | 0 | 27 | 0 | 80 | 107 |
| | Jul | 0 | 0 | 0 | 0 | 29 | 0 | 82 | 111 |
| | Aug | 1 | 0 | 0 | 0 | 26 | 0 | 80 | 107 |
| | Sep | 1 | 0 | 0 | 0 | 18 | 0 | 123 | 142 |
| | Oct | 0 | 0 | 0 | 0 | 17 | 1 | 59 | 77 |
| | Nov | 1 | 0 | 0 | 0 | 51 | 3 | 136 | 191 |
| | Dec | 0 | 0 | 0 | 0 | 63 | 72 | 146 | 281 |
| 2021 | Jan | 0 | 0 | 0 | 0 | 49 | 64 | 97 | 210 |
| | Feb | 0 | 0 | 0 | 2 | 48 | 11 | 88 | 149 |

*Rhinovirus testing started in February 2019

#### Legends to Supplementary Figures

**Supplementary figure 1**

Daily nationally reported new COVID-19 cases (al ages), January 2020 through February 2021, and cumulative number of reported cases, in relation to mitigation measures.

1. First case of COVID-19 identified in Israel, mandatory quarantine for all people entering Israel (02-21-2020)
2. Closure of all educational institutions; banning gathering of more than 10 people; closures of all malls, restaurants, hotels, clubs, fitness clubs, swimming pools, beaches, wedding halls (03-13-2020)
3. First nationwide lockdown (including during Passover, Ramadan and Independence day): required face masks when leaving home; closure of all educational facilities; movement and travel restrictions (limited within 100 meters from home); discontinuation of nonessential work and commerce; no public gathering; limited public transportation; closure of prayer services; closing of airport both for outgoing and incoming flights (04-08-2020)
4. First national lockdown relief period (phase 1): removal of the 100 meter limit on venturing from home; schools open to grade 1-3; reopening of street stores, malls and hair and beauty salons; allowing outdoor meetings not exceeding 20 people; weddings up to 50 attendees (05-01-2020)
5. First national lockdown relief period (phase 2); schools fully reopened; restaurants reopened; airport reopened, beaches and museums reopened; number of passengers in public transportation relaxed; houses of worship reopened for up to 50 people (05-17-2020)
6. End of school year (07-01-2020)
7. Beginning of ultra-Orthodox school year (08-21-2020)
8. Beginning of school year (09-01-2020)
9. Second lockdown (during the high holidays): closure of schools except for special education and boarding schools; no sitting in bars and restaurants (delivery service allowed); gathering limited to 10 people indoor and 20 people outdoor; closure of malls, stores (except food and pharmacies), hotels,

fitness clubs, swimming pools; outdoor prayer limited to 20 people, 10 people indoor; closure of airport (09-18-2020)

**Supplementary figure 2**

A- Dynamics of monthly nationwide IPD rates in children $<5$ years of age January 2016 through February 2021, and monthly newly nationally reported COVID-19 cases (all ages).

B- Incidence rate ratios (IRRs) of IPD in children $<5$ years of age by the period, January 2020 through February 2021 vs. 2016-2019.

**Supplementary figure 3**

A- Dynamics of monthly lower respiratory infections (LRIs) rates in children <5 years of age, January 2016 through February 2021, and month new nationally reported COVID-19 cases (all ages)

B- Incidence rate ratios (IRRs) of LRISs in children <5 years of age during the period January 2020-2021vs. January 2016 through February  2016-2019.

**Supplementary figure 4**

Monthly virus-positive nasal wash samples and CAAP rates in children <5 years of age, by specific virus, January 2016 through February 2021. Rhinovirus PCR testing was initiated only in February 2019.

min = Minimum

max = Maximum

**Supplementary figure 5**

Montly virus-positive nasal wash samples and NA-LRI rates in children <5 years of age, by specific virus, January 2016 through February 2021. Rhinovirus PCR testing was initiated only in February 2019.

min = Minimum

max = Maximum

**Supplementary figure 6**

A. Dynamics of monthly incidence rates of trauma visits, children <5 years of age  at the Soroka University Medical Center, January 2016 through February 2021, and monthly newly reported COVID-19 cases nationally, January 2020 through February 2021 (all ages).

B. Dynamics of monthly incidence rates of non-trauma visits, children <5 years of age, at the Soroka University Medical Center, January 2016 through February 2021, and monthly newly reported COVID-19 cases nationally, January 2020 through February 2021 (all ages).

min = Minimum

max = Maximum

**Supplementary figure 1**

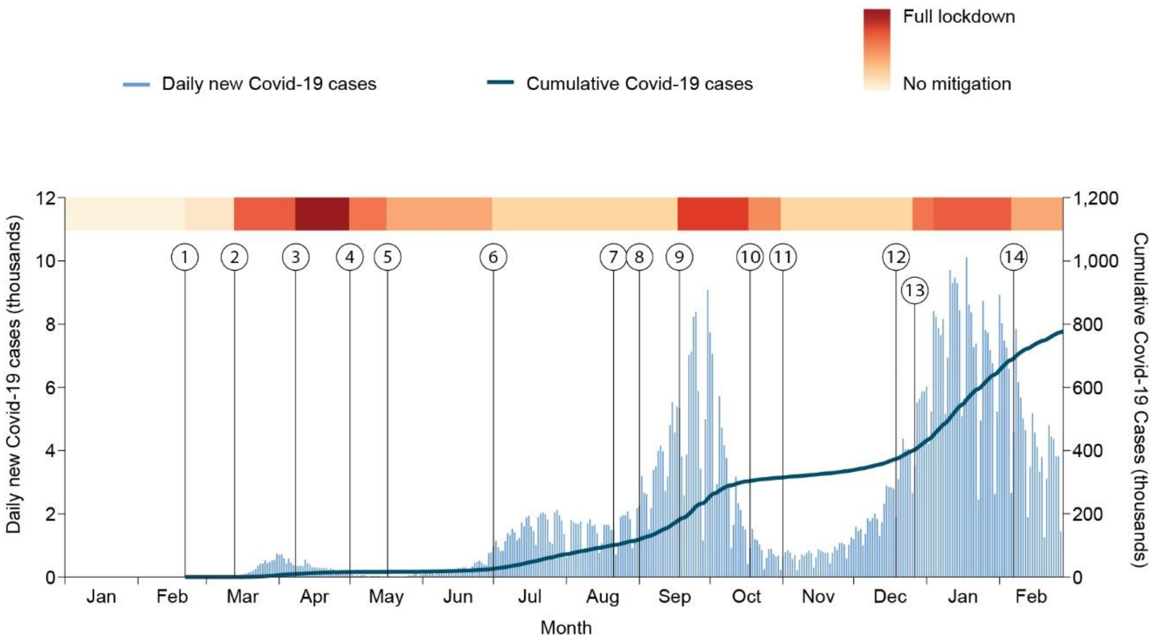

**Supplementary figure 2**

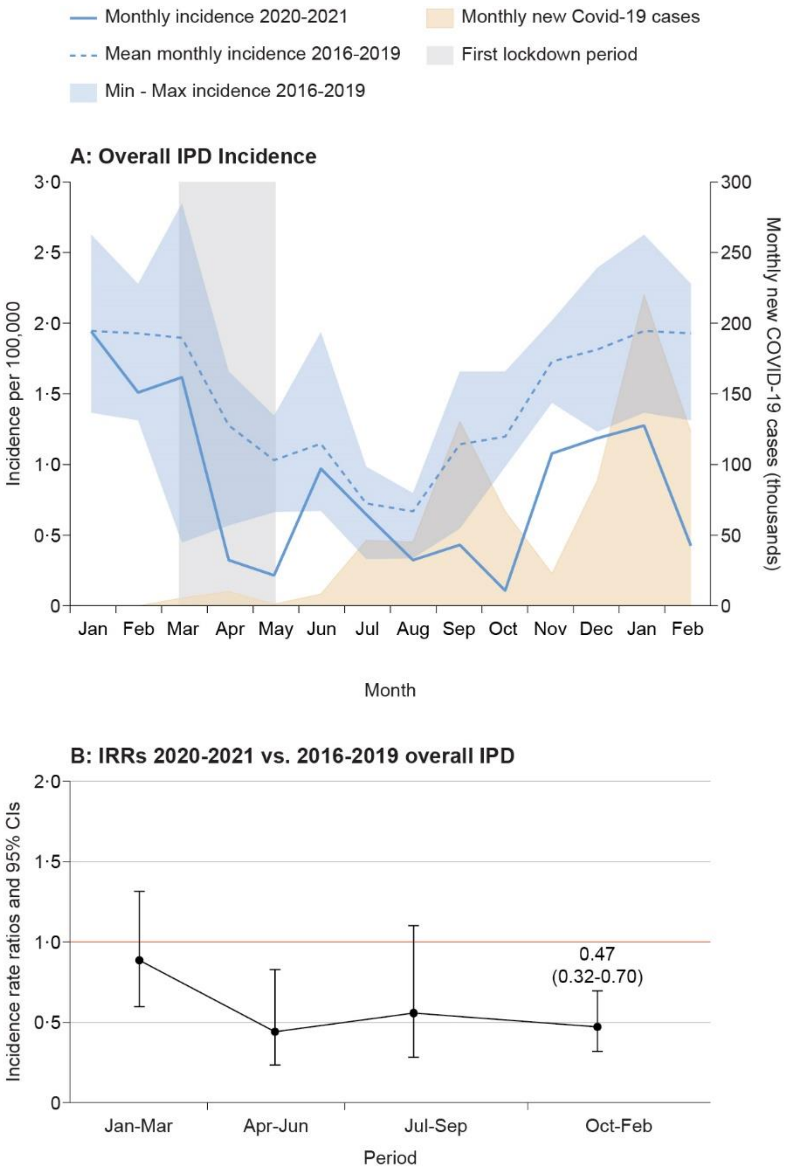

**Supplementary figure 3**

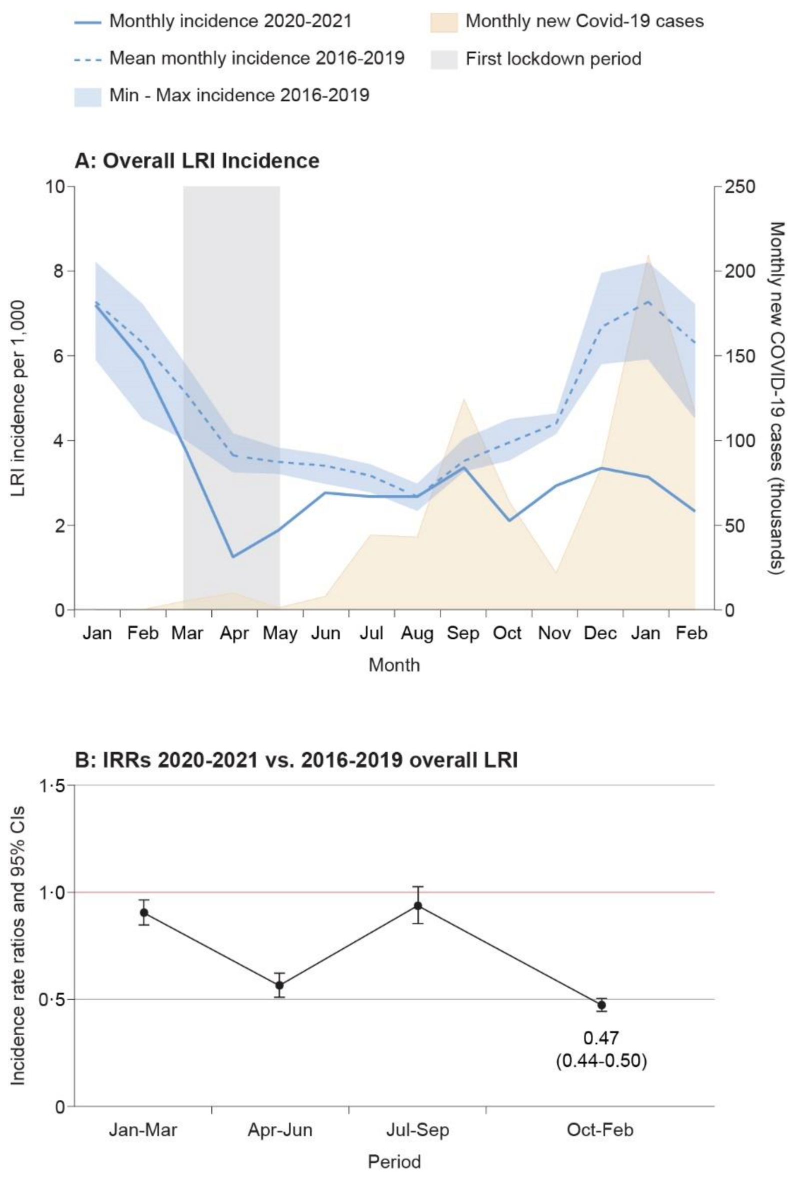

**Supplementary figure 4**

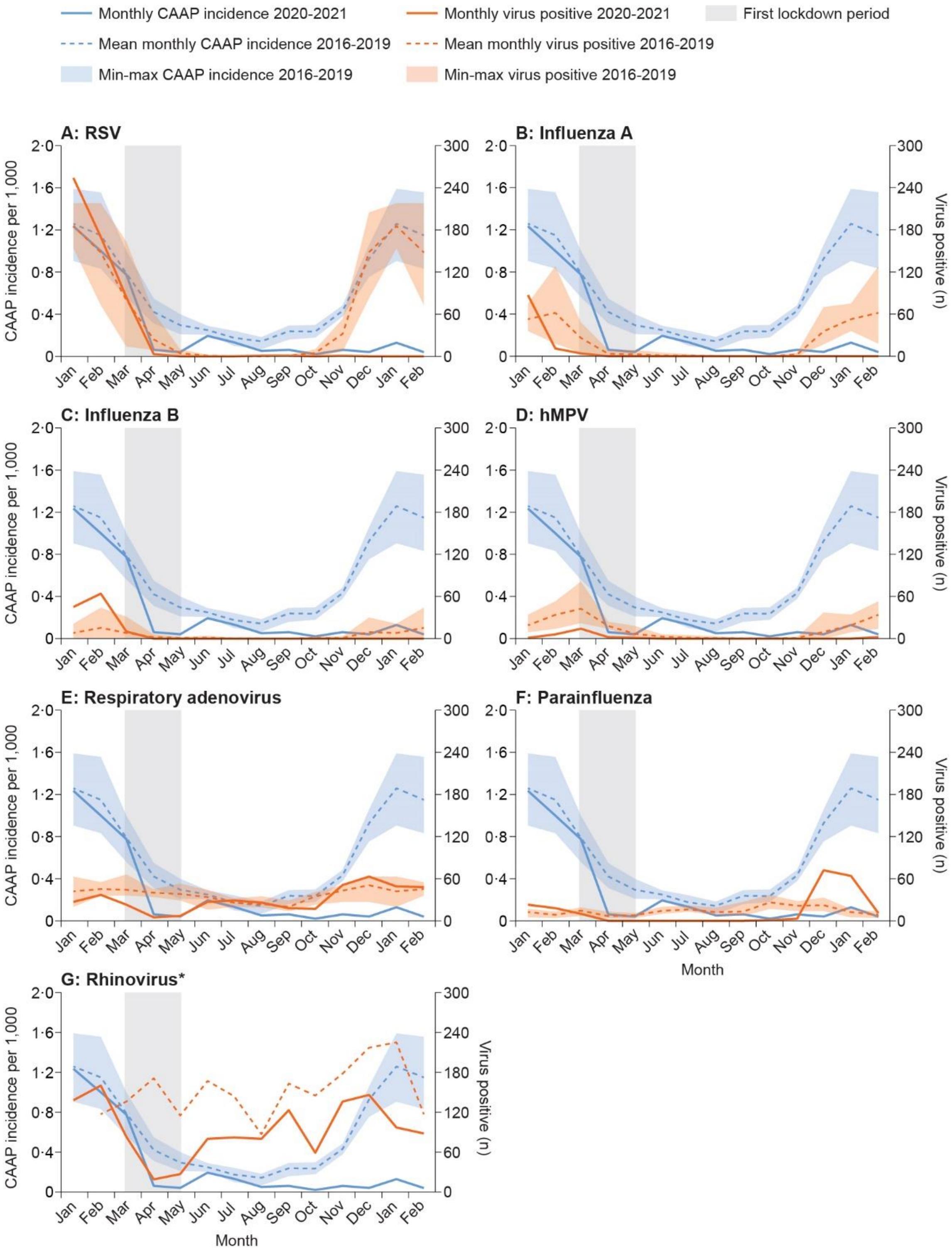

**Supplementary figure 5**

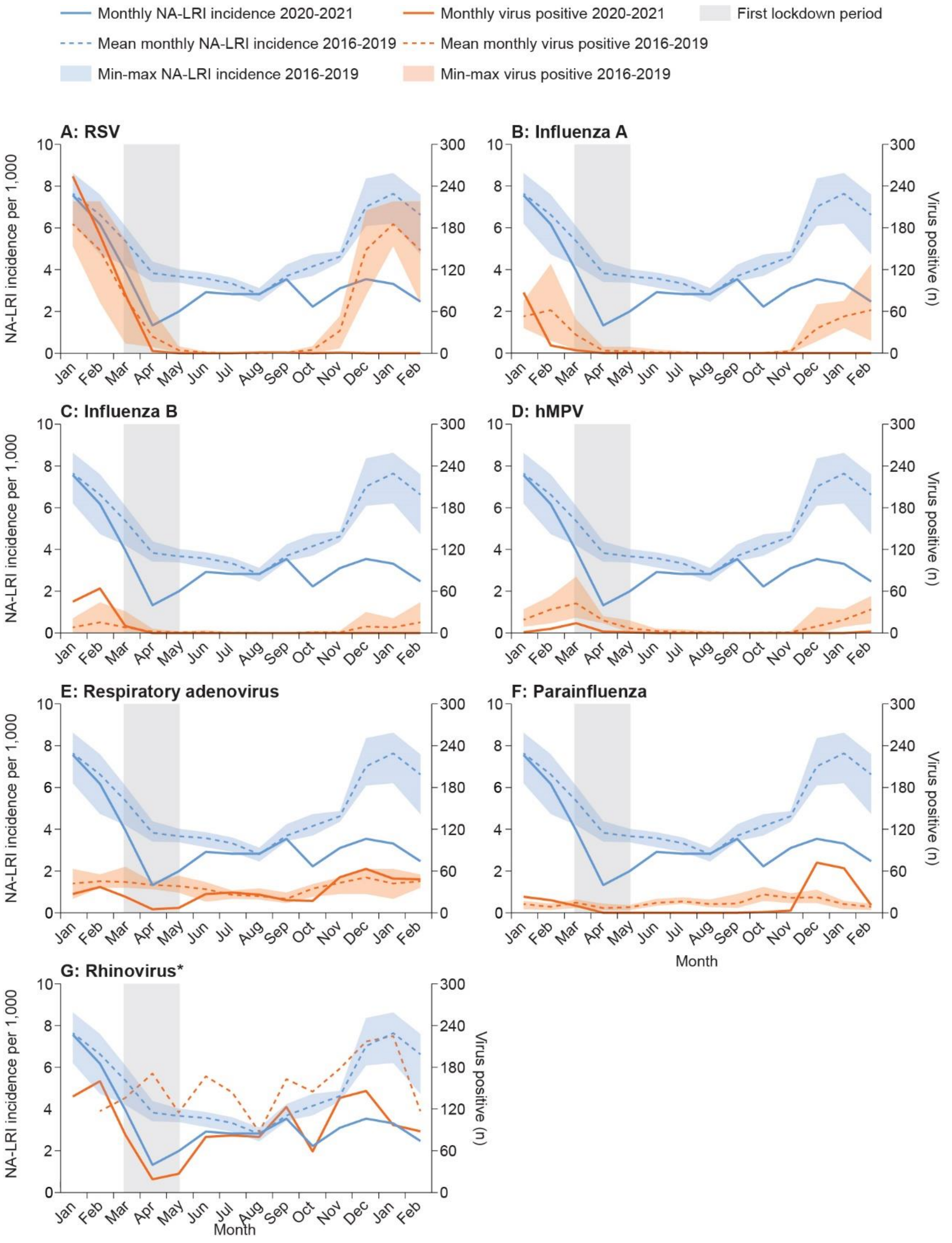

**Supplementary figure 6**

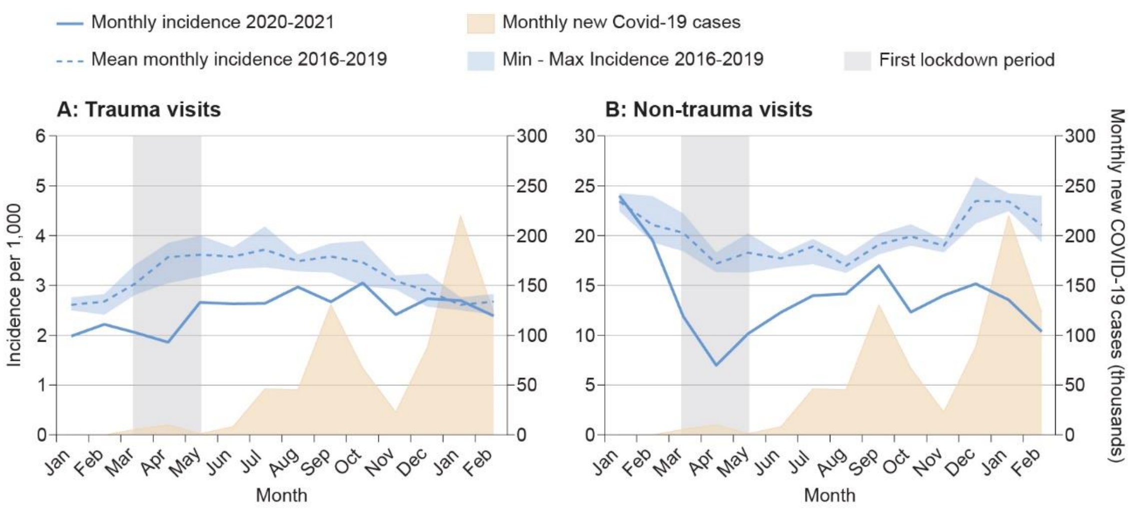
